# Evaluating Capabilities of Large Language Models: Performance of GPT4 on Surgical Knowledge Assessments

**DOI:** 10.1101/2023.07.16.23292743

**Authors:** Brendin R Beaulieu-Jones, Sahaj Shah, Margaret T Berrigan, Jayson S Marwaha, Shuo-Lun Lai, Gabriel A Brat

**Affiliations:** Department of Surgery, Beth Israel Deaconess Medical Center, Boston, MA; Department of Biomedical Informatics, Harvard Medical School, Boston, MA; Geisinger Commonwealth School of Medicine, Scranton, PA; Division of Colorectal Surgery, National Taiwan University Hospital, Taipei, Taiwan

**Keywords:** ChatGPT, artificial intelligence, language models, surgical education, surgery

## Abstract

**Background:** Artificial intelligence (AI) has the potential to dramatically alter healthcare by enhancing how we diagnosis and treat disease. One promising AI model is ChatGPT, a large general-purpose language model trained by OpenAI. The chat interface has shown robust, human-level performance on several professional and academic benchmarks. We sought to probe its performance and stability over time on surgical case questions.

**Methods:** We evaluated the performance of ChatGPT-4 on two surgical knowledge assessments: the Surgical Council on Resident Education (SCORE) and a second commonly used knowledge assessment, referred to as Data-B. Questions were entered in two formats: open-ended and multiple choice. ChatGPT output were assessed for accuracy and insights by surgeon evaluators. We categorized reasons for model errors and the stability of performance on repeat encounters.

**Results:** A total of 167 SCORE and 112 Data-B questions were presented to the ChatGPT interface. ChatGPT correctly answered 71% and 68% of multiple-choice SCORE and Data-B questions, respectively. For both open-ended and multiple-choice questions, approximately two-thirds of ChatGPT responses contained non-obvious insights. Common reasons for inaccurate responses included: inaccurate information in a complex question (n=16, 36.4%); inaccurate information in fact-based question (n=11, 25.0%); and accurate information with circumstantial discrepancy (n=6, 13.6%). Upon repeat query, the answer selected by ChatGPT varied for 36.4% of inaccurate questions; the response accuracy changed for 6/16 questions.

**Conclusion:** Consistent with prior findings, we demonstrate robust near or above human-level performance of ChatGPT within the surgical domain. Unique to this study, we demonstrate a substantial inconsistency in ChatGPT responses with repeat query. This finding warrants future consideration and presents an opportunity to further train these models to provide safe and consistent responses. Without mental and/or conceptual models, it is unclear whether language models such as ChatGPT would be able to safely assist clinicians in providing care.

## Background

Artificial intelligence (AI) models have the potential to dramatically alter healthcare by enhancing how we diagnosis and treat disease. These models could lead to increased efficiency, improved accuracy and personalized patient care. Successful healthcare-related applications have been widely reported.^1–10^ Within surgery, machine learning approaches that include natural language processing, computer vision, and reinforcement learning have each shown promise to advance care.^1, 11–14^ Still, despite the promise of AI to revolutionize healthcare, its use within the field is markedly limited compared to other industries. The severe implications of errors and empathy concerns regarding the use of AI in healthcare have led to cautious adoption.^7, 11, 15–18^

One promising recent model for use in healthcare is ChatGPT, a publicly-available, large language model trained by OpenAI.^19^ Released in November 2022, ChatGPT received unprecedented attention,^20^ given its notable performance across a range of medical and non-medical domains.^21^ ChatGPT has shown robust, human-level performance on several professional and academic benchmarks, including a simulated bar exam, the graduate record examination (GRE), numerous Advanced Placement (AP) examinations, and the Advanced Sommelier knowledge assessment.^19^ With regard to medical knowledge, an earlier version of ChatGPT was shown to perform at or near the passing threshold of 60% accuracy on the United States Medical Licensing Exam (USMLE).^22, 23^ In addition, ChatGPT has demonstrated robust performance on knowledge assessments in family medicine,^24^ neurosurgery,^25^ hepatology,^26^ and a combination of all major medical specialties.^27^ Moreover, ChatGPT has shown promise as a clinical decision support tool in radiology,^28^ pathology,^29^ and orthodontics.^30^ ChatGPT has also performed valuable clinical tasks,^31–34^ such as writing patient clinic letters, composing inpatient discharge summaries, suggesting cancer screening, and conveying patient education.^35^ Lastly, several studies have highlighted the potential impact of ChatGPT on medical education and research, with roles ranging from supporting nursing education to advancing data analysis and streamlining the writing of scientific publications.^32, 36–38^ The emergence of ChatGPT has reignited interest in exploring AI applications in healthcare; however, it has also provoked numerous concerns, regarding bias, reliability, privacy, and governance.^21, 26, 32, 36–41^

In the current study, we evaluate ChatGPT-4’s performance on two commonly used surgical knowledge assessments: The Surgical Council on Resident Education (SCORE) curriculum and a second case-based question bank for general surgery residents and practicing surgeons – which is referred to as Data-B and not identified due to copyright restrictions. SCORE is an educational resource and self-assessment used by many US residents throughout residency training.^42–45^ Data-B is principally designed for graduating surgical residents and fully-trained surgeons in preparation for the American Board of Surgery (ABS) Qualifying Exam (QE). These assessments were selected as their content represents the knowledge expected of surgical residents and board-certified surgeons, respectively. As such, it was thought that Form-B, while based on the same content area as SCORE, should include more higher-order management or multi-step reasoning questions, and that we may observe differential ChatGPT performance. The performance of ChatGPT on each of these assessments may provide important insights regarding ChatGPT-4’s capabilities at this point in time. Perhaps more importantly, in addition to assessing performance, this study investigates reasons for ChatGPT errors and assesses its performance on repeat queries. This latter objective represents a significant contribution to our current understanding of large language models – and a critical domain for research for safe and effective use of AI in healthcare.

## Methods

### Artificial Intelligence

ChatGPT (Open AI, San Francisco, CA) is a publicly-available, subscription-based AI chatbot that first launched in November 2022. It was initially derived from GPT-3 (Generative Pretrained Transformer) language models, which are pre-trained transformer models designed primarily to generate text via next word prediction. To improve performance for ChatGPT, initial GPT-3 models were further trained using a combination of supervised and reinforcement learning techniques.^50^ In particular, ChatGPT was trained using Reinforcement Learning from Human Feedback (RLHF), in which a reward model is trained from human feedback. To create a reward model, a dataset of comparison data was created, which was comprised of two or more model responses ranked by quality by a human AI trainer. This data could then be used to fine-tune the model using Proximal Policy Optimization.^46^

ChatGPT-PLUS is the latest development from Open-AI and employs GPT-4, which is the fourth iteration of the GPT family of language models.^19^ Details regarding the architecture and development of GPT-4 are not publicly available. It is generally accepted that GPT-4 was trained in a similar fashion as GPT-3 via RLHF. While specific technical details are unknown, Open AI states in their technical report that one of the main goals in developing GPT-4 was to improve the language model’s ability to understand and generate natural text, particularly in complex and nuanced scenarios. The report highlights improved performance of GPT-4, relative to GPT-3.5. For example, GPT-4 passed a simulated bar exam with a score in the 90^th^ percentile, whereas GPT-3.5 achieved a score in the 10^th^ percentile of exam takers. GPT-4 was officially released on March 13, 2023 and is currently available via the ChatGPT Plus paid subscription.

### Input Sources

Input questions were derived from two commonly used surgical educational resources:

1. SCORE: The Surgical Council on Resident Education (SCORE) is a nonprofit organization established in 2004 by the principal organizations involved in surgical education in the United States, including the American Board of Surgery (ABS) and the Association for Surgical Education (ASE). SCORE maintains a curriculum for surgical trainees, which includes didactic educational content and more than 2400 multiple-choice questions for self-assessment. A total of 175 self-assessment questions were obtained from the SCORE question bank. Access to the SCORE question bank was obtained through the research staff’s institutional access. SCORE was not part of the research team and did not participate in the study design and completion of research. Using existing functionality within SCORE, study questions were randomly selected from all topics, except systems-based practice; surgical professionalism and interpersonal communication education; ethical issues in clinical surgery; biostatistics and evaluation of evidence; and quality improvement. Fellowship-level questions were not excluded from study inclusion. Questions containing images were excluded from analysis. After exclusion, a total of 167 questions from SCORE were included in the study analysis.
2. Data-B: Data-B is an educational resource for practicing surgeons and senior surgical trainees, which includes case-based, multiple choice questions across a range of general surgical domains, including endocrine, vascular, abdomen, alimentary tract, breast, head and neck, oncology, perioperative care, surgical critical care, and skin/soft issue. A total of 120 questions were randomly selected for inclusion in the study. Questions containing images were excluded from analysis. After exclusion, 119 questions were included.

### Encoding

For input into ChatGPT, all selected questions were formatted two ways:

1. Open-ended (OE) prompting: Constructed by removing all answer choices and translating the existing question into an open-ended phrase. Examples include: “What is the best initial treatment for this patient?”; “For a patient with this diagnosis and risk factor, what is the most appropriate operative approach?”; or “What is the most appropriate initial diagnostic test to determine the cause of this patient’s symptoms?”
2. Multiple choice (MC) single answer without forced justification: Created by replicating the original SCORE or Data-B question verbatim. Examples include: “After appropriate cardiac workup, which of the following surgeries should be performed?”; “Which of the following laboratory values would most strongly suggest the presence of ischemic bowel in this patient?”; or “Which of the following options is the best next step in treatment?”

Open-ended prompts were deliberately varied to avoid systemic errors. For each entry, a new chat session was started in ChatGPT to avoid potential bias. To reiterate, all questions were inputted twice (once with open-ended prompting and once via the multiple choice format). Presenting questions to ChatGPT in two formats provided some insight regarding the capacity of a predictive language model to generate accurate, domain-specific responses without prompting.

### Assessment

Outputs from ChatGPT-4 were assessed for Accuracy, Internal Concordance and Insight by two surgical residents using the criteria outlined by Kung *et al.* in their related work on the performance of ChatGPT on the USMLE exam.^23^ Response accuracy (i.e., correctness) was assessed based on the provided solution and explanation by SCORE and Data-B, respectively. Internal response concordance refers to the internal validity and consistency of ChatGPT’s output—specifically whether the explanation affirms the answer and negates remaining choices without contradiction. Insight refers to text that is non-definitional, non-obvious and/or valid.^23^

Each reviewer adjudicated 100 SCORE questions and 70 Data-B questions, with 30% and 28% overlap, respectively. For overlapping questions, residents were blinded to each other’s assessment. Interrater agreement was evaluated by computing the Cohen kappa (κ) statistic for each question type (Supplemental Table 1).

For the combined set of 167 SCORE questions included in the study, the median performance for all human SCORE users was 65%, as reported in the SCORE dashboard. Reference data for Data-B is not available, preventing an exact comparison between ChatGPT and surgeon users.

In addition, we reviewed all inaccurate ChatGPT responses to multiple choice SCORE questions to determine and classify the reason for the incorrect output. The classification system for inaccurate responses was created by study personnel and designations were made by consensus. Reasons for inaccurate responses included: inaccurate information in complex question, inaccurate information in fact-based question; accurate information, circumstantial discrepancy; inability to differentiate relative importance of information; imprecise application of detailed information; and imprecise application of general information. A description of each error type, as well as representative examples are shown in **Table 1**.

**Table 1:**
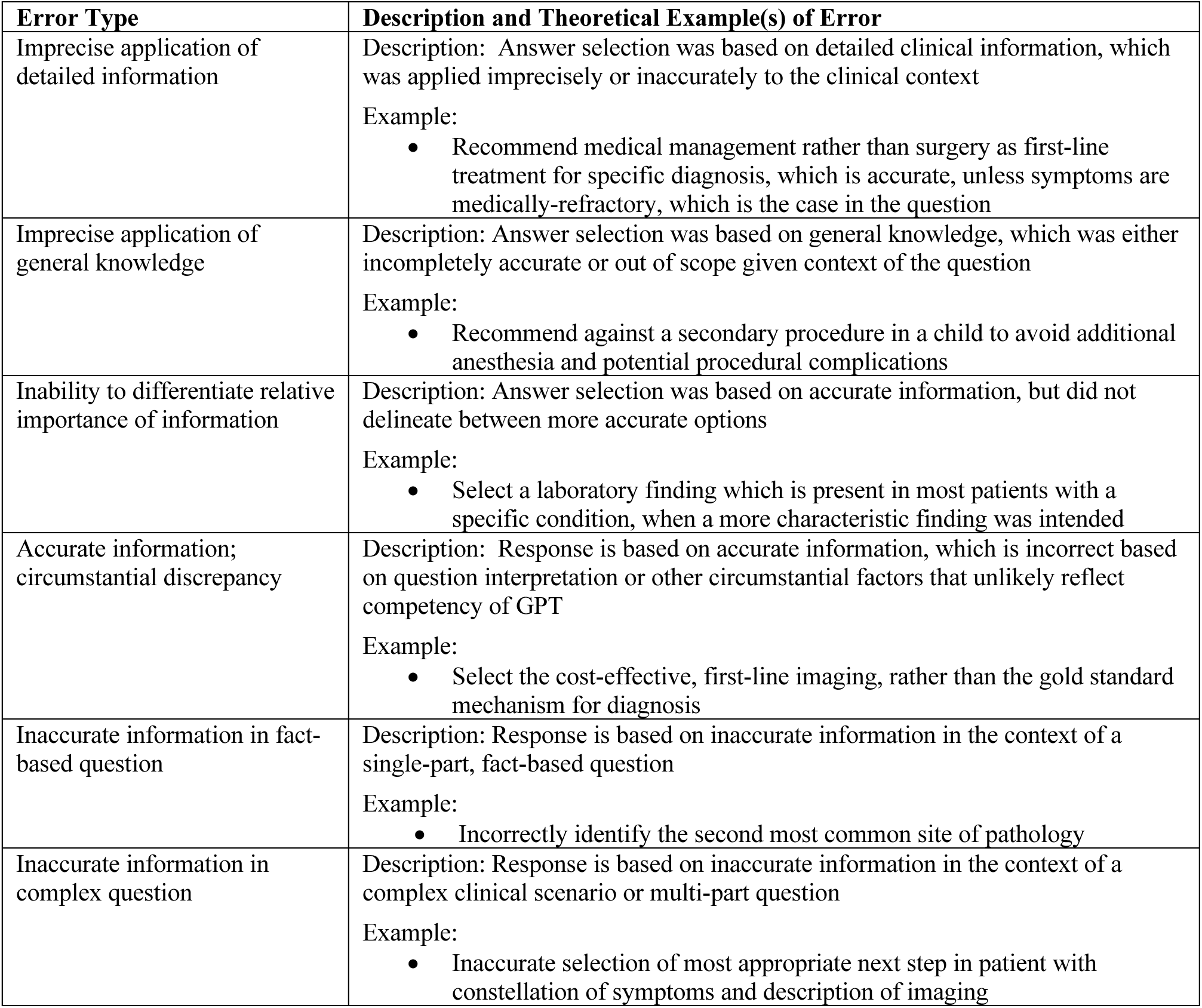
Classification of Error Type: Description and Examples.

To further assess the performance and reproducibility of GPT-4, all responses to SCORE questions (MC format) that were initially deemed inaccurate were re-queried. Second, ChatGPT responses were compared to the initial output to determine if the answer response changed and if it changed, whether the response was now accurate or if it remained inaccurate.

## Results

### Accuracy of ChatGPT Responses

A total of 167 SCORE and 112 Data-B questions were presented to ChatGPT. The accuracy of ChatGPT responses for OE and MC SCORE and Data-B questions is presented in **Figure 1**. ChatGPT correctly answered 71% and 68% of MC SCORE and Data-B questions, respectively. The proportion of accurate responses for OE questions was lower than for MC, particularly for SCORE questions, which is largely due to an increase in responses that were deemed indeterminate by study adjudicators in the setting of the open-ended format.

**Figure 1:**
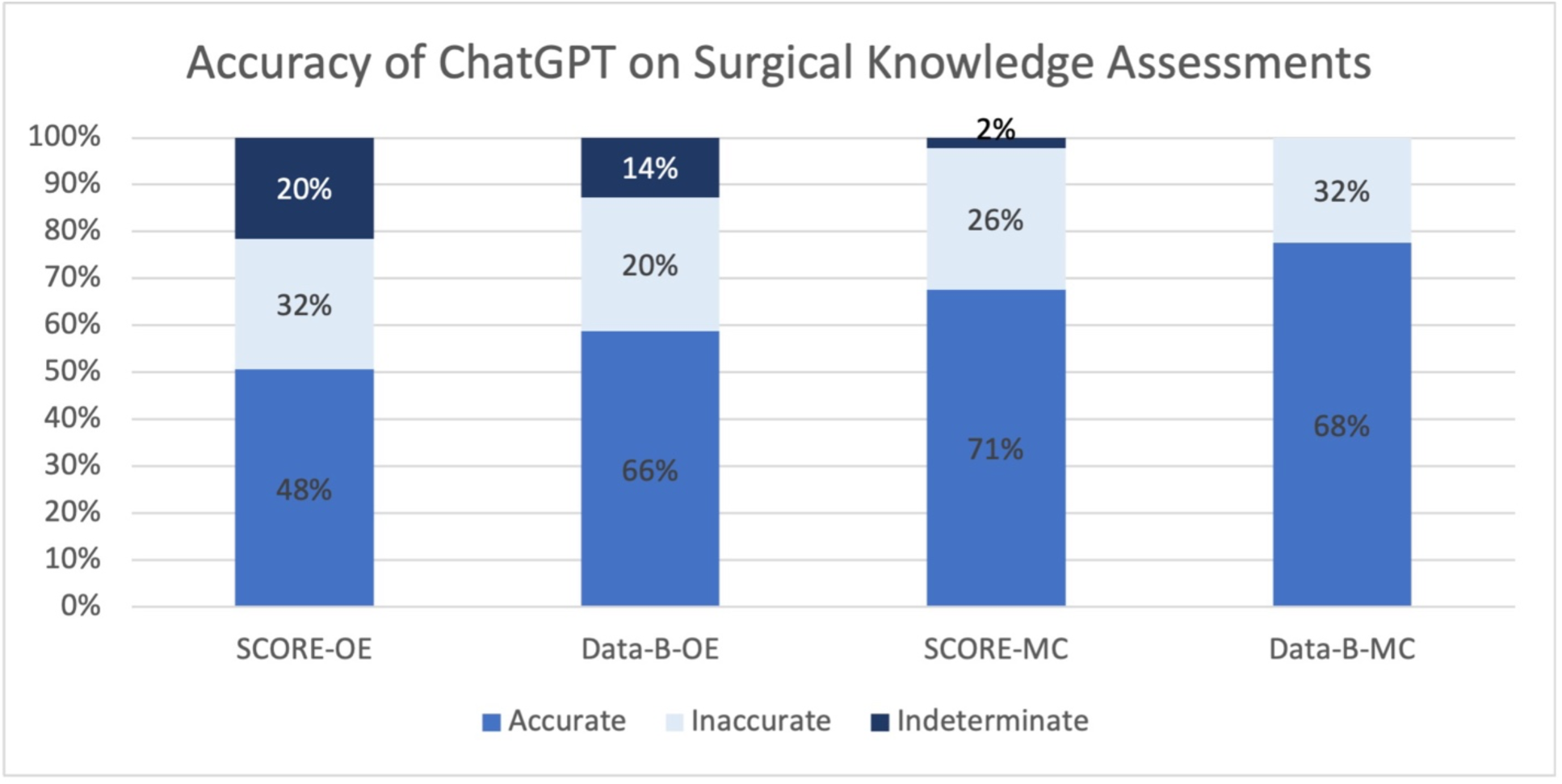
Accuracy of ChatGPT Output for Open-Ended and Multiple-Choice Questions. **Legend:** Surgical knowledge questions from SCORE and Data-B were presented to ChatGPT via two formats: open-ended (OE; left sided and multiple-choice (MC; right side). ChatGPT’s outputs were assessed for accuracy by surgeon evaluators. A total of 167 SCORE and 112 Data-B questions were presented to the ChatGPT interface. ChatGPT correctly answered 71% and 68% of multiple choice SCORE and Data-B questions, respectively.

### Internal Response Concordance of ChatGPT Responses

Internal Concordance was adjudicated by review of the entire ChatGPT response (**Table 2**). Overall internal response concordance was very high: 85.6% and 100% for OE SCORE and Data-B questions, respectively, and 88.6% and 97.3% for MC SCORE and Data-B questions. Among OE SCORE questions, internal response concordance was also assessed by accuracy subgroup (**Figure 2**). Concordance was nearly 100% (79/80) for accurate responses. Internally discordant responses were more frequently observed for inaccurate responses (33%, 31/75).

**Figure 2:**
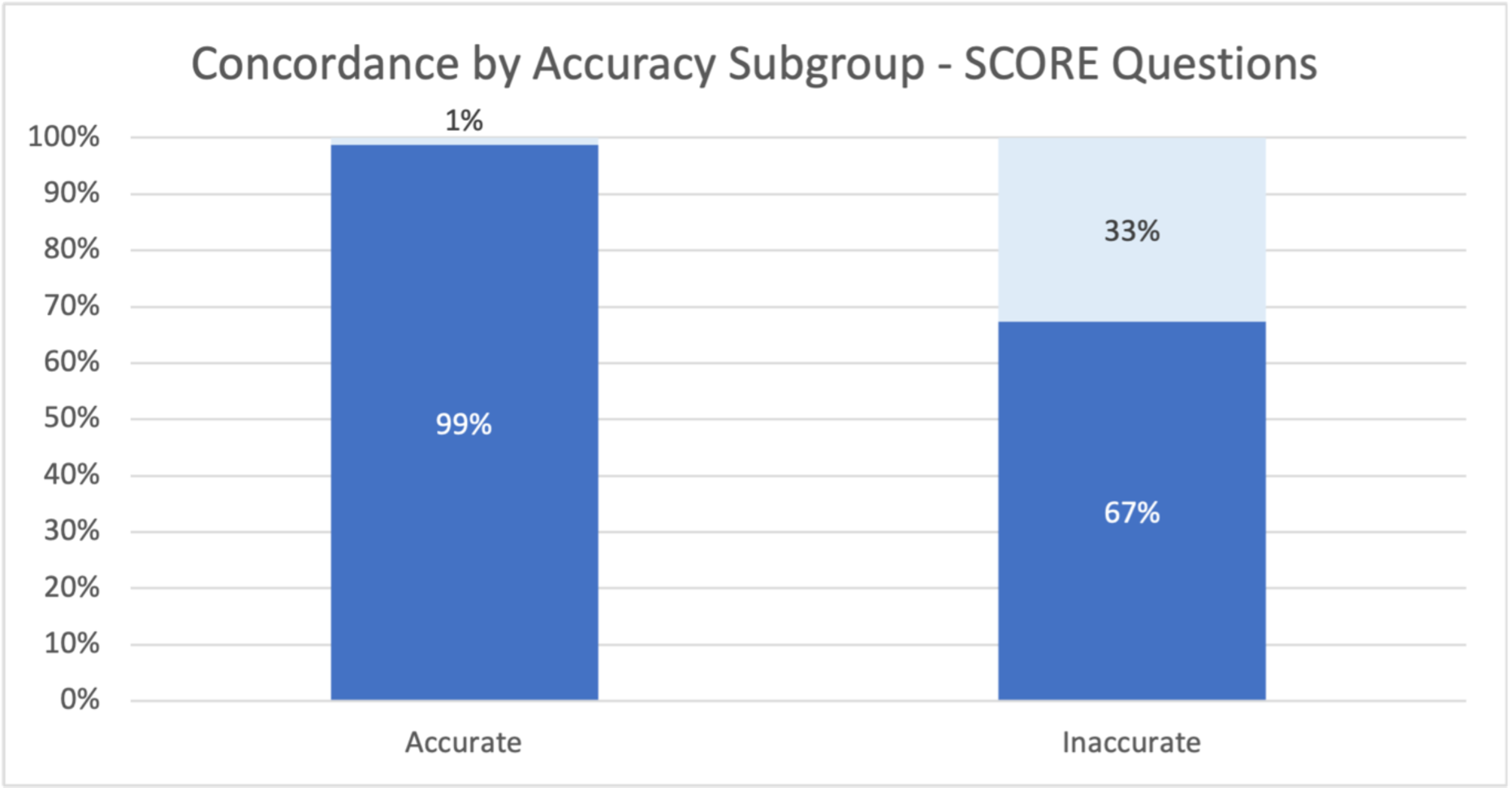
Internal Concordance by Accuracy Subgroup among SCORE Questions. **Legend:** SCORE questions were presented to ChatGPT via two formats: open-ended and multiple-choice. ChatGPT’s outputs to open-ended SCORE questions were assessed for internal concordance by accuracy subgroup. A total of 167 SCORE questions were presented to the ChatGPT interface. Concordance was nearly 100% (79/80) for accurate responses. Internally discordant responses were more frequently observed for inaccurate responses (33%, 31/75).

**Table 2:** Accuracy, Internal Concordance and Nonobvious Insights of ChatGPT Responses.

|  | Accuracy, N (%) | Internal Concordance | Insights |
| --- | --- | --- | --- |
| Open-Ended Format |  |  |  |
| SCORE | 80 (47.9%) | 143 (85.6%) | 111 (66.5%) |
| Data-B | 74 (66.1%) | 112 (100%) | 87 (77.7%) |
| Combined | 154 (55.2%) | 255 (91.4%) | 198 (71.0%) |
| Multiple Choice - Single Answer |  |  |  |
| SCORE | 119 (71.3%) | 148 (88.6%) | 106 (63.5%) |
| Data-B | 76 (67.9%) | 109 (97.3%) | 69 (61.6%) |
| Combined | 195 (69.9%) | 257 (92.1%) | 175 (62.7%) |

SCORE: Surgical Council on Resident Education;
Data-B: refers to a second commonly used surgical knowledge assessment and question bank

### Insights within ChatGPT Responses

For both OE and MC questions, approximately two-thirds of ChatGPT responses contained nonobvious insights (**Table 2**). Insights were more frequently observed for OE questions (SCORE: 66.5% versus 63.5%; Data-B: 77.7% versus 62.7%).

### Classification of Inaccurate ChatGPT Responses to MC SCORE Questions

Reasons for inaccurate responses are shown (**Table 3).** The most common reasons were: inaccurate information in a complex question (36.4%); inaccurate information in fact-based question (25.0%); and accurate information, circumstantial discrepancy (13.6%).

**Table 3:**
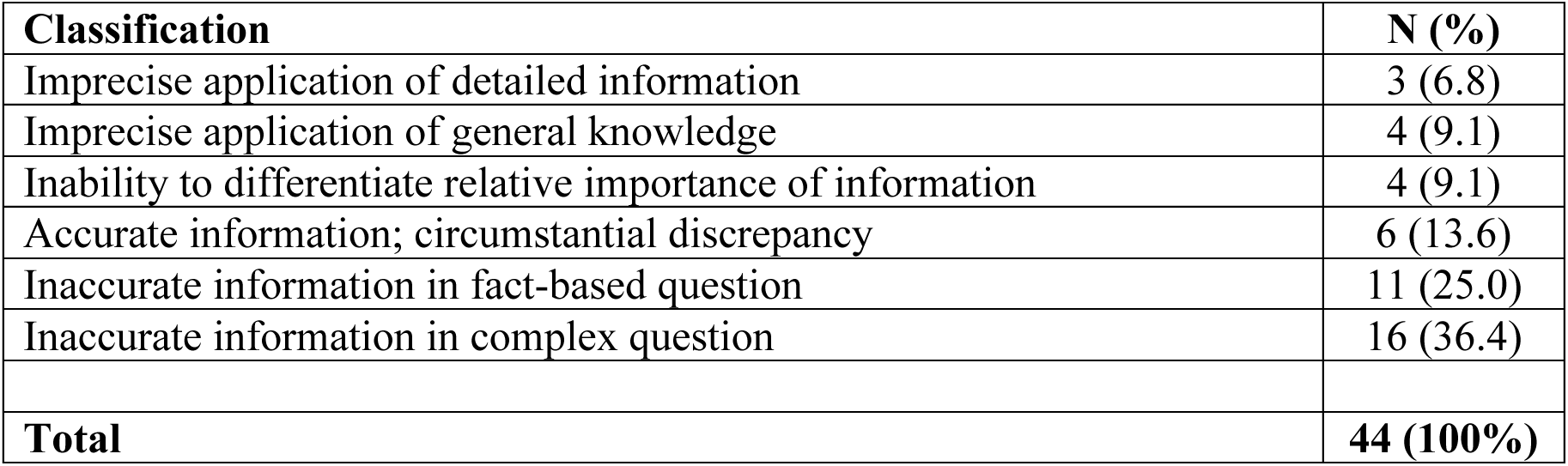
Classification of Inaccurate ChatGPT Responses for SCORE Questions (N=44)

| **Classification** | **N (%)** |
| --- | --- |
| Imprecise application of detailed information | 3 (6.8) |
| Imprecise application of general knowledge | 4 (9.1) |
| Inability to differentiate relative importance of information | 4 (9.1) |
| Accurate information; circumstantial discrepancy | 6 (13.6) |
| Inaccurate information in fact-based question | 11 (25.0) |
| Inaccurate information in complex question | 16 (36.4) |
| **Total** | **44 (100%)** |

**Table 4:** Outcome of Repeat Question for 44 Initially Inaccurate Responses to SCORE.

| **Outcome** | **N (%)** |
| --- | --- |
| No change in answer/response | 28 (63.6%) |
| Change in answer/response |  |
| Inaccurate to inaccurate | 10 (22.7%) |
| Inaccurate to accurate | 6 (13.6%) |
| **Total** | **44 (100%)** |

### Outcome of Repeat Question for Initially Inaccurate ChatGPT Responses

For all inaccurate ChatGPT responses to MC SCORE questions, the exact MC SCORE question was re-presented to the ChatGPT on a separate encounter, using a new chat. The accuracy of the response was assessed as prior. In total, the answer selected by ChatGPT varied between iterations for 16 questions (36.4% of inaccurate questions). The response remained inaccurate in 10/16 questions and was accurate on the second encounter for 6/16 questions. No change in the selected MC answer was observed in nearly two-thirds of cases (n=28, 63.6%).

## Discussion

To assess ChatGPT’s capabilities within the surgical domain, we assessed the performance of ChatGPT-4 on two surgical knowledge self-assessments. Consistent with prior findings in other domains, ChatGPT exhibited robust accuracy and internal concordance, near or above human-level performance. The study highlights the accuracy of ChatGPT within a highly specific and sophisticated field without specific training or fine-tuning in the domain. The findings also underscore some of the current limitations of AI including variable performance on the same task and unpredictable gaps in the model’s capabilities. In addition, the non-tiered performance of ChatGPT on SCORE and Data-B suggests a distinctiveness between human knowledge and/or learning and the development of language models. Nonetheless, the robust performance of a language model within the surgical domain – and potential to enhance its performance domain-specific training (i.e., high-yield surgical literature) – highlights its potential value to support and advance human tasks in clinical decision-making and healthcare. While human context and high-level conceptual models are needed for certain decisions and tasks within surgery, understanding the performance large language models will direct their future development such that AI tools are complementarily positioned within healthcare, offloading extraneous cognitive demands.

Foremost, within the surgical domain, ChatGPT demonstrated near or above human-level performance, with an accuracy of 71% and 68% on MC SCORE and Data-B questions, respectively. This is consistent with ChatGPT performance in other general and specific knowledge domains, including law, verbal reasoning, mathematics, and medicine.^19, 23^ The current findings are consistent with a study by Hopkins *et al.*, in which ChatGPT was tested on and achieved near human-level performance on a subset of questions from the Congress of Neurological Surgeons (CNS) Self-Assessment Neurosurgery (SANS).^25^

The current study utilizes two knowledge assessments, which are generally accepted to be tiered in difficulty, with SCORE principally designed for residents and Data-B targeted for senior residents and board-certified attending general surgeons. This design provides additional insight into the performance of ChatGPT relative to humans; we would anticipate that ChatGPT would perform superiorly on SCORE relative to Data-B. However, the near equivocal relative performance of ChatGPT on SCORE and Data-B suggests that its capabilities do not parallel those of surgical trainees. Learners progressively attain greater layers of context and understanding to expand their knowledge. A predictive language processing model such as ChatGPT does not improve in a similar manner, given the nature of its corpus of information and reinforcement-based training. It is an informal observation that SCORE questions often require more precise delineation of similar answer choices (e.g., distal pancreatectomy with splenectomy versus distal pancreatectomy alone), and Data-B generally requires a broader knowledge set to answer each question. The near equivocal performance suggests that a probabilistic algorithm like ChatGPT can function at a high level in both tasks, but it also highlights that the mental and conceptual models that providers use to develop their expertise should not be attributed to these models. Mental models have acknowledged limitations, but they allow physicians to think broadly during clinical encounters where information is limited. Importantly, such differences may lead to errors by the language model that experienced providers would consider basic or unlikely given the way we learn. Thus, it is still too early to assume language models can safely assist clinicians in providing care. Future research into how large language models perform, with specific attention to end-points beyond accuracy, is needed to direct further development and application of language models and related AI in surgery and healthcare.

Two additional findings warrant consideration. First, our analysis highlighted the kind of errors that ChatGPT makes on surgical knowledge questions. In 11 of 44 inaccurate responses (25%), the incorrect response related to a straightforward, fact-based query (e.g., What is the second most common location of blunt thoracic vascular injury after the aorta?). Second, we observed inconsistencies in ChatGPT responses. When erroneous responses were re-presented to the language model interface, output varied in one-third of instances, and responses were different and incorrect (e.g. select another multiple choice response) in two-thirds. These two findings highlight a substantial limitation of current predictive language models when the response changes over several days. Future performance metrics should include a measure of consistency as well as initial capability. Fine-tuning to the specific domain may improve the confidence of the model and subsequent consistency; this type of finding underscores the importance of implementing AI tools in a complementary fashion in healthcare, given the high costs of errors.

To our knowledge, this is the first study testing the performance of ChatGPT on knowledge assessments over multiple instances. The extraordinary results of a general-purpose model like ChatGPT highlight both the incredible opportunity that exists and the value of additional domain-specific fine tuning and reinforcement learning. In particular, future research is needed to assess ChatGPT’s performance within clinical encounters, rather than standardized knowledge assessments. Large language models such as ChatGPT lack a conceptual model, and this is fundamentally different from how humans diagnose and treat, and may be a major limitation of ChatGPT’s performance in clinical settings—as highlighted in a recent blog post by an emergency medicine physician who tested ChatGPT’s diagnostic capacity for a subset of recent clinical encounters.^52^ Without these mental or conceptual models, correct responses to deterministic questions, like the questions within SCORE and Data-B, do not necessarily imply that the model would be able to assist clinicians in providing care in its current form.

The current study has notable limitations. First, a relatively small bank of questions was used, which may not accurately reflect the broader surgical knowledge domain. Second, the assessment of accuracy and internal concordance for the open-ended responses may be biased, but we found significant inter-rater reliability to calm this concern. Third, and most importantly, it is possible that some of the questions and/or answers are available in some form online and may allow the model to draw on previous answers. While the content is easily accessible online, the specific questions are less likely to be available online as both SCORE and Data-B are not open-source assessments. Finally, a metric of human performance on Data-B is not readily available, though median performance is likely equivalent to SCORE, given the reported data on both the American Board of Surgery In-Training and Qualifying Examinations.

## Conclusion

Consistent with prior findings, the current study demonstrates the robust performance of ChatGPT within the surgical domain. Unique to this study, we demonstrate inconsistency in ChatGPT responses and answer selections upon repeat query. This finding warrants future consideration and demonstrates an opportunity for further research to develop tools for safe and reliable implementation in healthcare. Without mental models, it is unclear whether language models such as ChatGPT would be able to safely assist clinicians in providing care.

## Data Availability

All input to the ChatGPT interface and associated output were recorded. Due to copyright laws, this data is not presented in the current manuscript. However, pending requisite approval from the respective organizations, this data may be shared upon reasonable request.

## Supplemental Material

**Supplemental Table 1:** Interrater Agreement – Cohen kappa for OE and MC questions.

|  | Open-Ended Questions |  | Multiple Choice – Single Answer |  |
| --- | --- | --- | --- | --- |
|  | Cohen K | N | Cohen K | N |
| SCORE | 0.720 | 30 | 1.0 | 30 |
| Data-B | 0.681 | 20 | 1.0 | 20 |

SCORE: Surgical Council on Resident Education
Data-B: refers to a second commonly used surgical knowledge assessment and question bank

